# Prediction of atrial fibrillation and stroke using machine learning models in UK Biobank

**DOI:** 10.1101/2022.10.28.22281669

**Authors:** A. Papadopoulou, D. Harding, G. Slabaugh, E. Marouli, P. Deloukas

## Abstract

We employed machine learning (ML) approaches to evaluate 2,199 clinical features and disease phenotypes available in the UK Biobank as predictors for Atrial Fibrillation (AF) risk. After quality control, 99 features were selected for analysis in 21,279 prospective AF cases and equal number of controls. Different ML methods were employed, including LightGBM, XGBoost, Random Forest (RF), Deep Neural Network (DNN),) and Logistic Regression with L1 penalty (LR). In order to eliminate the black box character of the tree-based ML models, we employed Shapley-values (SHAP), which are used to estimate the contribution of each feature to AF prediction. The area-under-the-roc-curve (AUROC) values and the 95% confidence intervals (CI) per model were: 0.729 (0.719, 0.738) for LightGBM, 0.728 (0.718, 0.737) for XGBoost, 0.716 (0.706,0.725) for DNN, 0.715 (0.706, 0.725) for RF and 0.622 (0.612, 0.633) for LR. Considering the running time, memory and stability of each algorithm, LightGBM was the best performing among those examined. DeLongs test showed that there is statistically significant difference in the AUROCs between penalised LR and the other ML models. Among the top important features identified for LightGBM, using SHAP analysis, are the genetic risk score (GRS) of AF and age at recruitment. As expected, the AF GRS had a positive impact on the model output, i.e. a higher AF GRS increased AF risk. Similarly, age at recruitment also had a positive impact increasing AF risk. Secondary analysis was performed for the individuals who developed ischemic stroke after AF diagnosis, employing 129 features in 3,150 prospective cases of people who developed ischemic stroke after AF, and equal number of controls in UK Biobank. The AUC values and the 95% CI per model were: 0.631 (0.604, 0.657) for XGBoost, 0.620 (0.593, 0.647) for LightGBM, 0.599 (0.573, 0.625) for RF, 0.599 (0.572, 0.624) for SVM, 0.589 (0.562, 0.615) for DNN and 0.563 (0.536, 0.591) for penalised LR. DeLongs test showed that there is no evidence for significant difference in the AUROCs between XGBoost and all other examined ML models but the penalised LR model (pvalue=2.00 E-02). Using SHAP analysis for XGBoost, among the top important features are age at recruitment and glycated haemoglobin. DeLongs test showed that there is evidence for statistically significant difference between XGBoost and the current clinical tool for ischemic stroke prediction in AF patients, CHA2DS2-VASc (pvalue=2.20E-06), which has AUROC and 95% CI of 0.611 (0.585, 0.638).

## Introduction

Atrial fibrillation (AF) is the most common cardiac arrythmia, which is characterised by a rapid and irregular heartbeat [1-5]. The incidence of AF is increasing rapidly with 12.1 million people expected to be affected by 2030 [6]. This is mainly attributed to the ageing of the population, along with changes in lifestyle [5, 7, 8]. AF, besides doubling the risk of cardiovascular mortality, is associated with increased risk of stroke, ischemic heart disease, heart failure and cognitive dysfunction [1, 4, 8, 9]. More specifically, AF quintuple the risk for ischemic stroke, independent of age [6, 10]. Additionally, the contribution of AF to ischemic stroke increases exponentially with age; a 1.5% attribution at 50-59 years reaches 23.5% for the age range 80-89 [6]. However, AF is sometimes asymptomatic, and thus remains undetected [3, 5], and subsequently the ischemic stroke risk attributed to AF is under-estimated [6, 11].

In recent years, machine learning (ML) algorithms have gained popularity in the field of medicine and specifically in disease prediction, classification of medical images and diagnosis. ML models use a hypothesis-free approach; there are no prior assumptions either among the input features or between the features and the outcome. Thus, it is possible to reveal new features, along with non-linear associations amongst them, which would have not been discovered using traditional statistical models. ML models have been proven to improve the accuracy of disease prediction, although they have a “black box” character and a different way of interpretating results than the traditional models [12-15].

There have been several studies that employed ML methods for prediction of circulatory diseases. A recent study by Raghunathan et al. [16] employed Deep Neural Networks (DNN) in 430,000 patients recorded in Geisinger’s clinical MUSE database between 1984 to 2019 with no history of AF, within 1-year of an ECG, and reported a model for AF prediction with an area under the receiver operating characteristic (AUROC) of 0.85. They also reported that 62% of patients who had a stroke caused by AF within 3 years of an ECG, with no prior AF diagnosis, would have been identified by their prediction tool before the stroke occurred. Another study by Su et al. [17] employed four ML models to predict modified Rankin Scale (mRS) at hospital discharge and in-hospital deterioration for acute ischemic stroke patients enrolled on the Stroke Registry in Chang Gung Healthcare System (SRICHS). Random Forest (RF) performed well in both outcomes; the AUROC was 0.829 for discharge mRS and 0.710 for in-hospital deterioration.

The aim of this study is to develop ML models for prediction of: 1) AF and 2) ischemic stroke in patients with AF, using UK-Biobank’s real-world clinical data, questionnaires, hospital episode statistics data and genomic data. To achieve this, five types of ML models, including extreme gradient boosting (XGBoost) [18], light gradient boosting machine (LightGBM) [19], RF [20], support vector machine (SVM) [21], DNN [22], and penalised logistic regression (LR) [23] as a baseline model, were constructed, and their predictive performances were compared. Besides the comparison of the model performances, we also focused on the ranking of feature importance by employing the SHapley Additive exPlanations (SHAP) [24], in order to unravel each feature’s contribution both to AF and to ischemic stroke prediction in AF patients.

## Methods

### Overview of the research framework

We included clinical data, phenotypes, lifestyle, and medications from the UKB. We imputed the missing values and employed a feature selection process, described in more detail at *Data pre-processing* section, in order to reduce the number of features employed to the ones relative to the outcome. Then six models, including XGBoost, LightGBM, RF, SVM, DNN and penalised LR were used to create the predictive models. The model’s hyperparameters were optimised using 10-fold cross validation at the training dataset, which was the 80% of the original one. The ML models were validated on the test dataset and their performances were compared. Lastly, we employed the SHAP explanations to reveal the features’ contributions to the prediction.

### Data pre-processing

We examined the UKB, a prospective cohort of 502,492 participants, aged 37-73 years old, recruited between 2006 and 2010. The dataset includes blood measurements, clinical assessments, anthropometry, cognitive function, hearing, arterial stiffness, hand grip strength, sociodemographic factors, lifestyle, family history, psychosocial factors and dietary intake [25]. Related individuals were removed, and the remaining dataset for analysis included 454,118 participants. Furthermore, we incorporated medications as features, derived from field 20003 (https://biobank.ndph.ox.ac.uk/showcase/field.cgi?id=20003). Additionally, clinical data were employed, coded in ICD10, derived from the Hospital Episodes Statistics (HES), which are linked to the UKB. From these, we constructed phenotype codes or “phecodes”, using a phecode map [26], which are aggregated ICD10 codes defining specific diseases or traits. We employed only the umbrella phecode categories. Detailed list of all the features that we examined can be found at the *Supplementary_Material.xlsx (Table_S1, Table_S2, Table_S3)*. Moreover, we created two polygenic scores (PGS) which were included as features for the prediction of ischemic stroke in people with AF. The first one is the AF score, based on 94 genome-wide variants derived from the Roseli et al. [27] genome-wide association study (GWAS) for AF. The second is the Ischemic STROKE score, based on 28 genome-wide variants derived from the Malik et al. [28] GWAS for ischemic stroke. The AF SCORE was also employed as a feature both for the prediction of AF and for the ischemic stroke in AF patients.

The investigator phenotypes dataset from UKB includes 2,199 fields for 454,118 participants. We set answers “Do not know” and “Prefer not to answer” as NA and removed features that had more than 25% missingness, resulting in 390 investigator phenotypes. Afterwards, we imputed the missing values using a multivariate imputer that estimates each feature from all the others, using *IterativeImputer* from Python [29]. Then, we added 419 phecodes, available for 278,177 participants, derived from HES in UKB. Lastly, we added the medications from field 20003 (https://biobank.ndph.ox.ac.uk/showcase/field.cgi?id=20003), after applying one-hot-encoding, resulting in 1,289 medications for 294,698 participants.

Next, we decided to balance the outcome sample size, since imbalanced data has a negative impact on ML procedures [30]. The classification algorithms have the tendency to get biased estimates towards the majority class, ignoring the minority class. This happens because most of the classifying methods aim to maximize the accuracy rate, meaning the number of correctly classified observations [31, 32]. Therefore, we employed the *fixed under-sampling* technique from Python [33], which is a process for reducing the number of samples in the majority class; the control group in our case. The algorithm randomly selects samples from the control group, in order to have equal representation of both classes. After balancing the outcome, we used *VarianceThreshold* from Python [29], which eliminates all features whose variance does not meet a threshold of 90%. Additionally, we removed the continuous correlated fields using Pearson correlation, at a 0.8 threshold; features strongly correlated with the outcome were maintained. Next, we performed feature selection in order to reduce the computational cost via dimensionality reduction [34], achieve higher classification accuracy by eliminating the noise, and include the most relevant features for the disease prediction. A recent paper by Ramos-Pérez et al. [35], suggests that the best practice is for the fixed under-sampling technique to precede the feature selection. Therefore, we filtered all the remaining features using recursive feature elimination with cross-validation from Python [29] in order to find the optimal number of features to include in the ML models.

### Outcome-AF

We removed participants from the UKB that had cardiac dysrhythmias before the time of enrolment, with one or more of the following codes: non-cancer illness code, self-reported (1471, 1483); operation code (1524); diagnoses – main/secondary ICD10 (I44, I44.1-I44.7, I45, I45.0-I45.6, I45.8-I45.9, I46, I46.2, I46.8-I46.9, I47, I47.0-I47.2, I47.9, I48, I48.0-4, I48.9, I49, I49.0-I49.5, I49.8-I49.9, R00.0, R00.1, R00.2, R94.3, Z86.7, Z95.0, Z95.8-Z95.9); underlying (primary/secondary) cause of death: ICD10 (I44, I44.1-I44.7, I45, I45.0-I45.6, I45.8-I45.9, I46, I46.2, I46.8-I46.9, I47, I47.0-I47.2, I47.9, I48, I48.0-4, I48.9, I49, I49.0-I49.5, I49.8-I49.9, I60-I61, I63-I64 (NOT I63.6), R00.0, R00.1, R00.2, R94.3, Z86.7, Z95.0, Z95.8-Z95.9); diagnoses – main/secondary ICD9 (4273, 430, 431, 4339, 4340, 4341, 4349, 436); operative procedures – main/secondary OPCS (K57.1, K62.1-4). In total, 20,584 participants were excluded, having at least one of the above conditions, before enrolment in the UKB.

AF cases were defined when having one or more of the following codes: non-cancer illness code, self-reported (1471, 1483); operation code (1524); diagnoses – main/secondary ICD10 (I48, I48.0-4, I48.9); underlying (primary/secondary) cause of death: ICD10 (I48, I48.0-4, I48.9); operative procedures – main/secondary OPCS (K57.1, K62.1-4). In total, 21,279 people developed one of the conditions described above, after enrolment in the UKB.

Based on the data pre-processing described above, 21,279 prospective AF cases and equal number of controls, along with 99 features were included in the ML models (*Supplementary_Material.xlsx, Table_S4*). Table 1 includes the baseline characteristics of the examined participants.

**Table 1:** Baseline characteristics for the 21,279 prospective AF cases and equal number of controls.

|  | **Total** | **AF cases** | **AF controls** |
| --- | --- | --- | --- |
| **Sex** |  |  |  |
| Females | 20231 (47.5%) | 8122 (38.2%) | 12109 (56.9%) |
| Males | 22327 (52.5%) | 13157 (61.8%) | 9170 (43.1%) |
| **Age (mean, sd)** | 59 (8) | 62 (6) | 57 (8) |
| **Ethnicity** |  |  |  |
| EUR | 41042 (96.9%) | 20791 (97.7%) | 20251 (95.0%) |
| AFR | 535 (1.2%) | 154 (0.7%) | 381 (1.8%) |
| EAS | 127 (0.3%) | 31 (0.2%) | 96 (0.5%) |
| SAS | 854 (1.6%) | 303 (1.4%) | 551 (2.7%) |
| **Comorbidities** |  |  |  |
| Diabetes | 6434 (15.1%) | 4423 (20.8%) | 2011 (9.5%) |
| Hypertension | 22019 (51.7%) | 14810 (69.6%) | 7209 (33.9%) |
| Smoking |  |  |  |
| Never | 23273 (54.7%) | 11627 (54.6%) | 11646 (54.7%) |
| Previous | 14791 (34.8%) | 7389 (34.7%) | 7402 (34.8%) |
| Current | 4494 (10.6%) | 2263 (10.6%) | 2231 (10.5%) |
| Cholesterol lowering medication | 7459 (17.5%) | 3712 (17.4%) | 3747 (17.6%) |
| **History of heart diseases** | 21102 (49.6%) | 11233 (52.8%) | 9869 (46.4%) |
| **History of stroke** | 12317 (28.9%) | 6581 (30.9%) | 5736 (26.9%) |

### Outcome-AF & Stroke

Cases were defined as participants who developed ischemic stroke after AF diagnosis in the UKB with one or more of the following codes: diagnoses – main/secondary ICD10 (I63, I63.0-9, I64); diagnoses – main/secondary ICD9 (434, 436); underlying (primary/secondary) cause of death: ICD10 (I63, I63.0-9, I64). Thus, 3,150 people developed ischemic stroke after AF diagnosis and were included as cases, and the controls were people diagnosed with AF and did not develop stroke, as far as the data allow us to know.

Based on the data pre-processing described above, 3,150 prospective cases who developed stroke after AF diagnosis and equal number of controls were included, along with 129 features were included in the ML models (Supplementary_Material.xlsx, Table_S7).

### Machine learning models

#### XGBoost

XGBoost is a “scalable machine learning system for tree boosting” [18]. This machine learning technique handles sparse data, incorporating a novel tree learning algorithm, runs ten times faster than similar algorithms, using parallel and distributed computing, and employs out-of-core computation, allowing the manipulation of massive datasets [18, 36]. In more detail, XGBoost uses regression trees in a sequential learning process as weak learners into a single strong model, where each tree attempts to correct the residuals in the predictions made by previous trees. Regression trees include a continuous score on each leaf, which is the last node once the tree has grown. For a specific observation, the algorithm uses the decision rules in the trees to classify it into the leaves. The sum of the scores on each leaf is the final prediction [15, 18, 37].

#### LightGBM

Machine learning methods relying on Gradient Boosting Decision Tree (GBDT) scan all the data instances, for all the features, in order to calculate the information gain for each possible split. As a result, the computational time and complexity will increase as the features accumulate. To this end, LightGBM [19] is introduced, which is an improved and lighter version of XGBoost. There are two techniques incorporated at LightGBM algorithm that contribute towards this improvement. Firstly, in the Gradient-based One-Side Sampling (GOSS) technique, instances that have larger gradients contribute more to the information gain, and the instances with smaller gradients are randomly sampled to provide accurate and fast estimation. Secondly, the Exclusive Feature Bundling (EFB) technique reduces the number of effective features. For datasets that are sparse, many features are mutually exclusive; they will rarely take nonzero values at the same time. Therefore, to reduce the dataset’s dimensions, such features are tied into one, reducing complexity of the algorithm [17, 19, 38, 39].

#### Random Forests (RF)

Random forest is a popular ensemble learning method using bootstrap aggregation (bagging) and feature randomness, in order to build an uncorrelated forest of several decision trees [20]. At the bagging step, each one of the decision trees is constructed from a random sample, drawn with replacement, from the training set. Once the tree is built, then a random subset of features is employed to split each node, which results in low correlation among the decision trees. Afterwards, the final prediction of RF is the result of the majority voting of all decision trees, leading to more accurate results [12, 17, 40].

#### Deep Neural Networks (DNN)

Deep learning is a subdomain of ML attempting to learn many levels of representation using multiple layers. These layers transform the data in a non-linear way, and as a result, more complex structure and relationships can be discovered. This method is inspired by the human brain, using a series of connected layers of neurons that constitute a whole network, including at least three layers: input, hidden and output. The input layer consists of multiple neurons, which use as input the original features. Then, the hidden layers can be more than one, depending on the complexity of the dataset. Each layer includes multiple nodes, and each node from the previous layer is connected to each one from the next layer, constituting a fully connected or dense network. Lastly, the output layer, using a sigmoid activation function, concludes in a number between 0 and 1, which represents the probability belonging to one of the two classes [22, 41-44].

#### Support Vector Machine (SVM)

Support Vector Machine (SVM) is a high accuracy ML model, which can deal with non-linear spaces. The basic idea is to map the input data into a higher dimension feature space, using a kernel function, which can be either linear, polynomial, radial basis function (rbf) or sigmoid. Then, a linear decision surface is created to classify the outcome, with properties that satisfy the generalisation of the algorithm. The linear decision surface is more commonly called hyperplane; the optimal one classifies the data by using the maximal margin of the hyperplane, employing a small percentage of the training data, which are named support vectors. It is supported that if the optimal hyperplane is created from a few support vectors, then the algorithm can be generalised, even in a space with infinite dimensions [21, 45, 46]. The SVM model is not easily interpretable, however it is included in our study in order to compare the predictive performance with the rest of the ML models [47].

#### Logistic regression-L1 penalty

One of the most common and easily interpretable models is the logistic regression, which is used to predict the outcome when it is classified in one of two classes. The Least Absolute Shrinkage and Selection Operator (Lasso) [48] - L1 regularisation - efficiently reduces the number of features of large datasets, and it has been proven to produce optimal sparse estimates when the true vector is sparse. To achieve this, it shrinks the coefficients of correlated and redundant features to zero. This method performs shrinkage and automatic feature selection in parallel. L1 regularisation has been proven to be effective when selection of relevant features, among plenty irrelevant ones, needs to be conducted [49, 50].

#### Cross-validation

The ML model aims to optimise the general model performance on datasets different from the ones used to train them [44]. Therefore, evaluating the generalisation of ML methods requires the data to be split in three non-overlapping sets of training/validation/test, using grid search, combined with stratified 10-fold cross-validation (CV), maintaining the same proportion of cases and controls in each fold. Grid search is performed using 9 sets for the parameter tuning, and the 1 remaining set is used for validation. This process is repeated 10 times, until every set is used once for training and once for validation. The best parameters for the model correspond to the highest score, which is calculated by averaging the results from all repetitions. The test dataset is used to check for overfitting and unbiased evaluation of the final model [29, 41, 42]. Tables with the hyperparameter values that were examined for each model can be found in *Supplementary_Material.xlsx*.

#### SHAP

ML models, although accurate and capable of capturing the non-linear relationships, are complex to interpret. A more widespread method for interpretation is SHapley Additive exPlanations (SHAP) [32, 37]. This is important, since ML models until recently were treated as “black boxes” [51]. We want to understand each feature’s contribution to the prediction, by calculating their explanations, using cooperative game theoretic tools [40, 52].

The SHAP values are in theory the best solution up to now, however time-consuming, since all possible combinations need to be calculated. TreeExplainer is an expansion of SHAP, employing tree nodes instead of linear models for the estimation of Shapley values. The Shapley values of a tree-based algorithm are calculated as the weighted average of the Shapley values corresponding to individual trees. Thus, it is commonly used to explain tree-based machine learning models, such as random forests and gradient boosted trees, reducing tremendously the computation time. In parallel, consistency and local accuracy are preserved [51, 53, 54].

Additionally, SHAP values seem to overcome the interpretability issue by employing both global and local interpretation for analysis methods that use trees. Global explanation relies on the effect of input features on the whole model, and local interpretation depicts the effect of input features on single predictions [24, 51, 53].

For the methods presented above, Python programming language was employed [55].

## Results

### AF

We examined 21,279 prospective AF cases and an equal number of controls in UKB including 99 features (*Supplementary_Material.xlsx (Table_S4*)) and using five ML models to predict AF. The results of the ML models presented in this section correspond to the optimal hyperparameters, derived after 10-fold cross-validation from the examined values included in *Supplementary_Material.xlsx* (*Table_S5*). SVM did not converge after running 10 days and utilising 16 cores in Queen Mary’s Apocrita HPC facility^1^.

For the test dataset, Table 2 summarises AUROC, accuracy, precision, recall and F1 score for each model. The best AUROC value was achieved with LightGBM (0.79; Table 2) albeit De-Long’s test (Table 3) showed that there is no evidence for significant difference in the AUROCs between LightGBM and XGBoost, DNN, or RF. In contrast, DeLong’s test showed that there was statistically significant difference in the AUROCs between LightGBM and penalised LR (pvalue=1.38E-02), after considering multiple correction. Actually, the AUROC of penalised LR differed from the AUROC of all other examined ML models based on DeLong’s test and this was statistically significant (Table 3). The AUROC curves for the 5 ML models in the test dataset are shown in Figure 1.

**Table 2:**
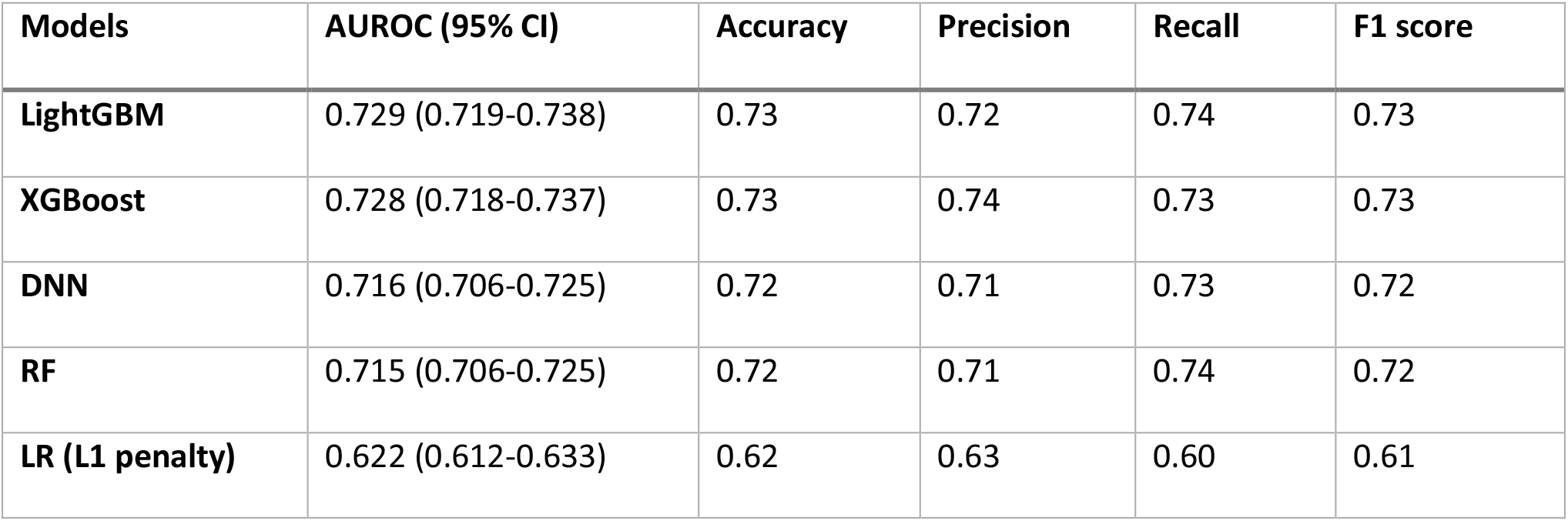
Performance of the ML models for AF prediction, on the test dataset, under various metrics.

| Models | AUROC (95% CI) | Accuracy | Precision | Recall | F1 score |
| --- | --- | --- | --- | --- | --- |
| **LightGBM** | 0.729 (0.719-0.738) | 0.73 | 0.72 | 0.74 | 0.73 |
| **XGBoost** | 0.728 (0.718-0.737) | 0.73 | 0.74 | 0.73 | 0.73 |
| **DNN** | 0.716 (0.706-0.725) | 0.72 | 0.71 | 0.73 | 0.72 |
| **RF** | 0.715 (0.706-0.725) | 0.72 | 0.71 | 0.74 | 0.72 |
| **LR (L1 penalty)** | 0.622 (0.612-0.633) | 0.62 | 0.63 | 0.60 | 0.61 |

**Table 3:** DeLong’s test for the ML model comparisons for AF prediction.

| Models | LightGBM | XGBoost | DNN | RF |
| --- | --- | --- | --- | --- |
| **LightGBM** | - |  |  |  |
| **XGBoost** | 8.28E-01 | - |  |  |
| **DNN** | 3.67E-02 | 5.78E-02 | - |  |
| **RF** | 1.17E-02 | 2.44E-02 | 9.91E-01 | - |
| **LR (L1 penalty)** | 1.38E-32 | 8.82E-32 | 2.41E-24 | 5.73E-27 |

<sup>1</sup> This research utilised Queen Mary's Apocrita HPC facility, supported by QMUL Research-IT. http://doi.org/10.5281/zenodo.438045

**Figure 1:**
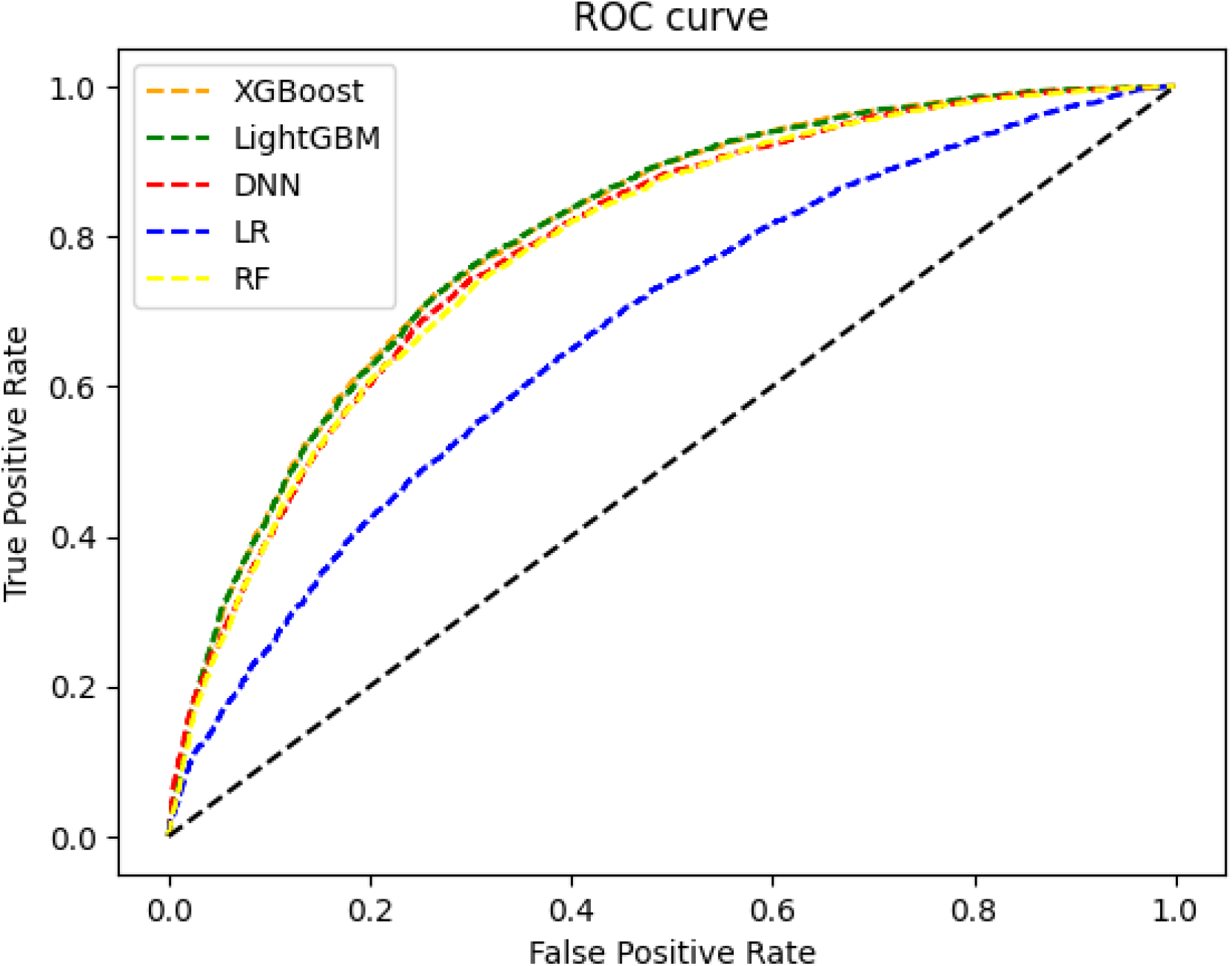
AUROC for each ML model for AF prediction in the test dataset.

To estimate the contribution of each feature in each of the 5 models we assessed for prediction of AF, we employed the TreeExplainer SHAP analysis, which is accurate, fast and stable (see Methods). Figure 2 displays the top 20 features, ranked according to their SHAP value, for the LightGBM model; features are listed in descending order, starting with the most significant for AF prediction. SHAP values depict the distribution of the effect of each feature on the model output.

**Figure 2:**
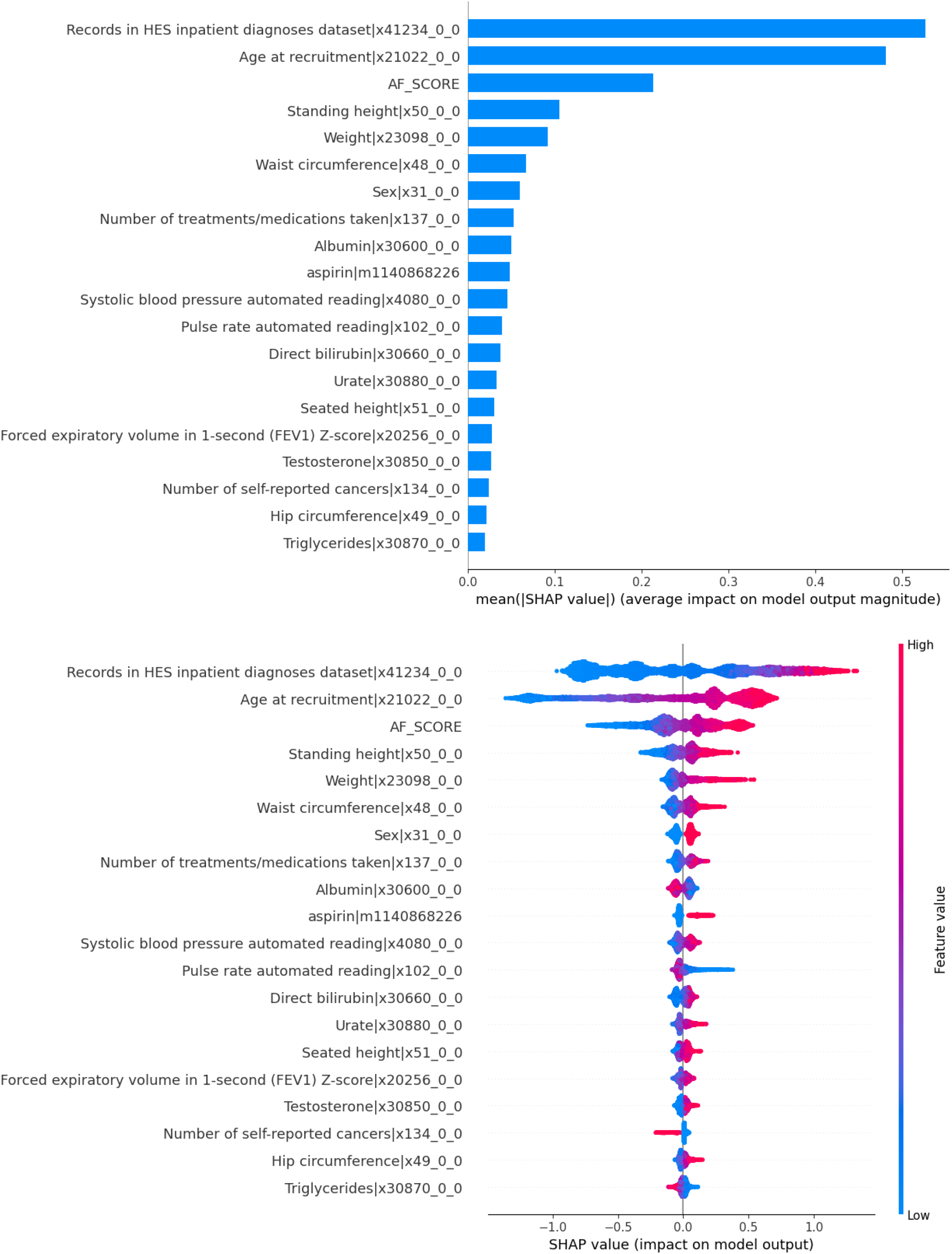
In both plots the top 20 features are depicted, in descending order, for the AF prediction on the test dataset, employing LightGBM model. On the top is the feature importance plot of the mean absolute SHAP values (x-axis) for the top 20 features (y-axis). On the bottom is the summary plot of the SHAP values (x-axis) for the top 20 features (y-axis), showing the distribution of the impact that each feature has on the model. Each dot represents a participant. The red dots represent a high feature value and blue dots represent a low feature value for each participant. For example, the AF SCORE had a positive impact on the model output, i.e., a higher AF SCORE increased AF risk.

Based on Figure 2, SHAP analysis reveals that the top 3 most important variables contributing to the model were “Records in HES inpatient diagnoses dataset” (fieldID 41234), “Age at recruitment” (fieldID 21022) and “AF SCORE”, using the unweighted sum of increasing alleles from Roseli et al. [27]. All the features’ contributions, based on SHAP analysis, can be found in *Supplementary_Material.xlsx (Table_S6)*.

### AF & Stroke

We examined 3,150 prospective cases who developed ischemic stroke after being diagnosed with AF, and an equal number of controls in UKB including 129 features (*Supplementary_Material.xlsx (Table_S7)* and using six ML models to predict ischemic stroke in AF cases. As indicated previously results correspond to the optimal hyperparameters (*Supplementary_Material.xlsx (Table_S8))*.

For the test dataset, Table 4 summarises the AUROC, accuracy, precision, recall and F1 score for each of the six models assessed for prediction of ischemic stroke in AF cases. The best AUROC value was achieved for XGBoost (Table 4). DeLong’s test (Table 5) showed that there is no evidence for significant difference in the AUROCs between XGBoost and all other examined ML models but the penalised LR model (pvalue=2.00 E-02).

**Table 4:**
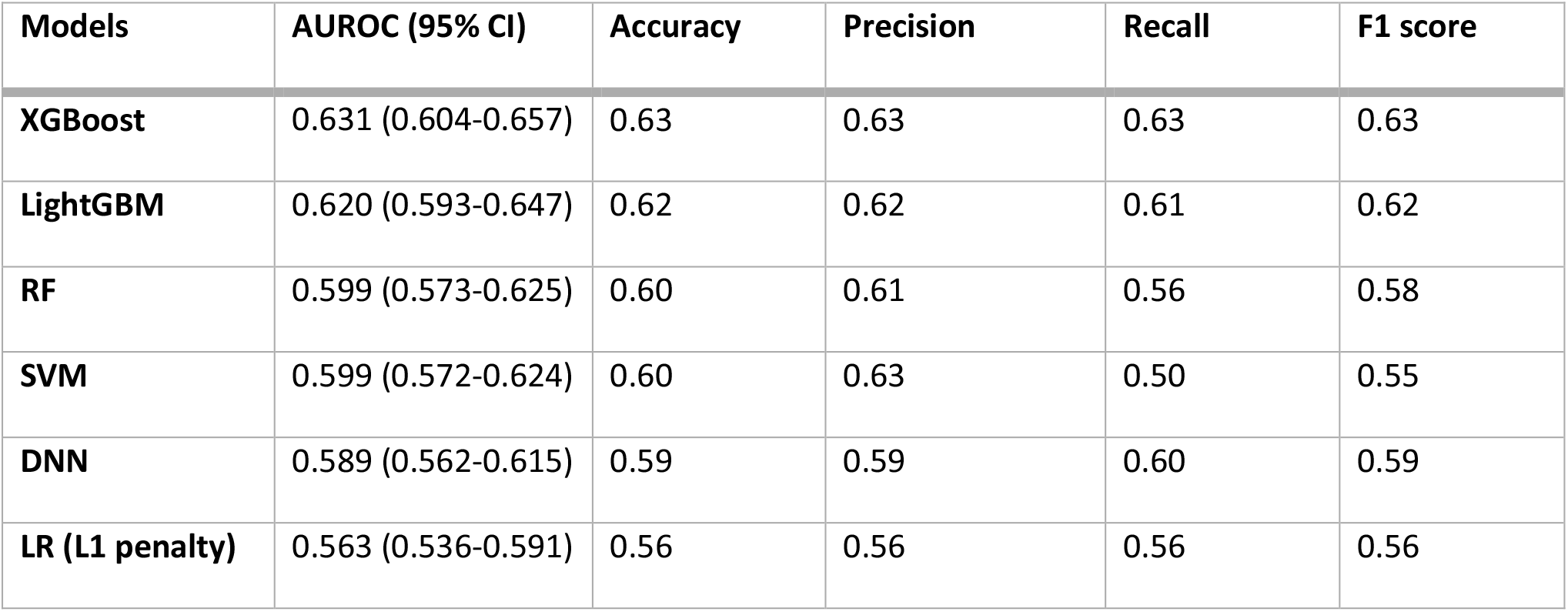
Performance of the ML models for the prediction of ischemic stroke in AF patients, on the test dataset, under various metrics.

**Table 5:** DeLong’s test for the ML model comparisons for ischemic stroke prediction in AF patients.

| Models | XGBoost | LightGBM | RF | SVM | DNN |
| --- | --- | --- | --- | --- | --- |
| **XGBoost** | - |  |  |  |  |
| **LightGBM** | 5.65E-01 | - |  |  |  |
| **RF** | 1.33E-01 | 3.45E-01 | - |  |  |
| **SVM** | 1.71E-01 | 3.75E-01 | 9.80E-01 | - |  |
| **DNN** | 1.34E-01 | 2.89E-01 | 7.54E-01 | 7.45E-01 | - |
| **LR (L1 penalty)** | 2.00E-02 | 5.70E-02 | 2.56E-01 | 4.50E-01 | 2.54E-01 |

As shown in Figure 4, SHAP analysis revealed that the 3 most important variables contributing to prediction of ischemic stroke in AF cases in the model were “Records in HES inpatient diagnoses dataset” (fieldID 41234), “Age at recruitment” (fieldID 21022), and “Glycated haemoglobin (HbA1c)” (fieldID 30750). *Supplementary_Material.xlsx (Table_S9)* lists the contribution of each of the 129 features in the model based on SHAP analysis.

**Figure 3:**
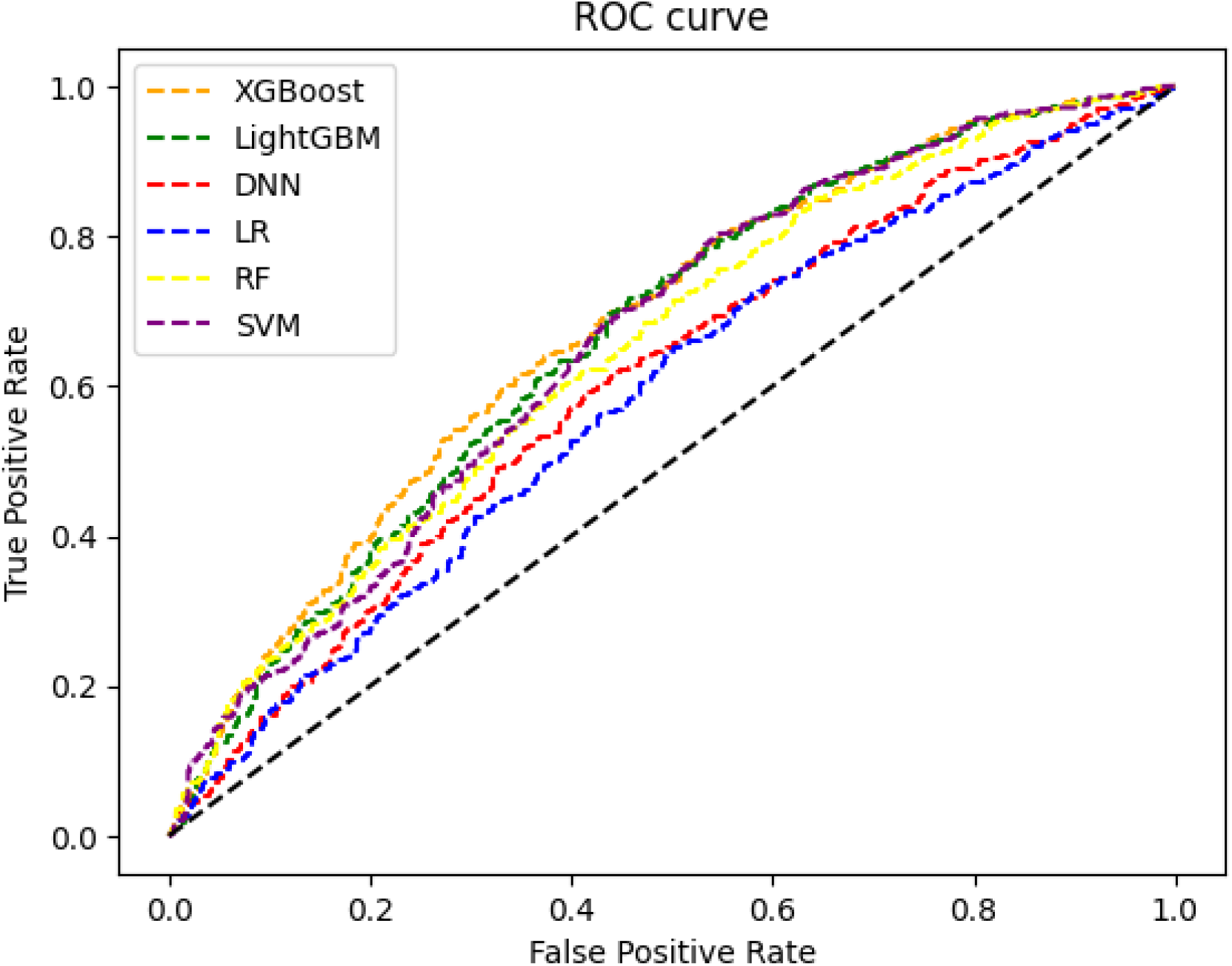
AUROC for each ML model for predicting the development of ischemic stroke in AF patients, on the test dataset.

**Figure 4:**
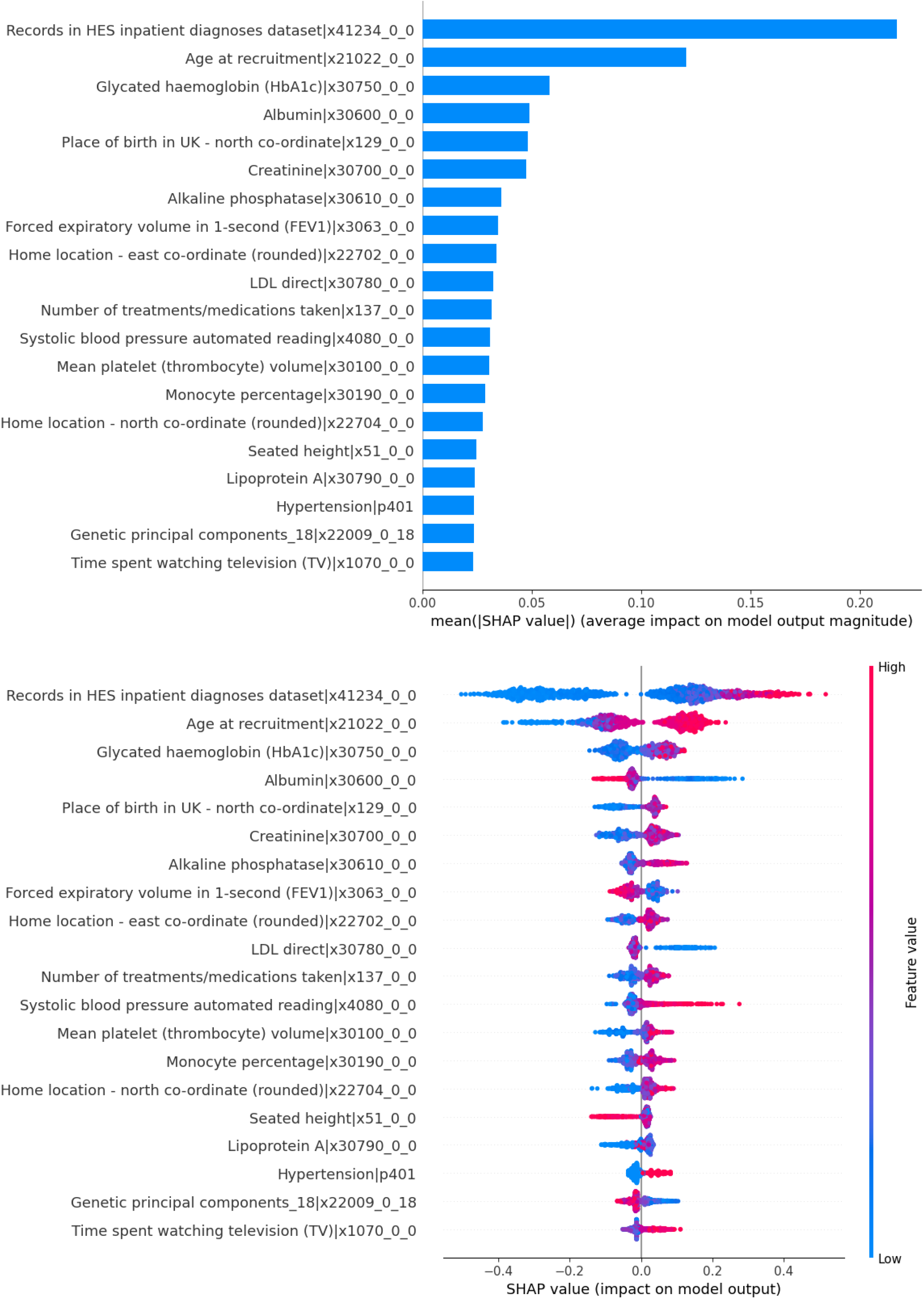
In both plots the top 20 features are depicted, in descending order, for the development of ischemic stroke in AF patients, on the test dataset, employing XGBoost model. On the top is the feature importance plot of the mean absolute SHAP values (x-axis) for the top 20 features (y-axis). On the bottom is the summary plot of the SHAP values (x-axis) for the top 20 features (y-axis), showing the distribution of the impact that each feature has on the model. Each dot represents a participant. The red dots represent a high feature value and blue dots represent a low feature value for each participant.

### Comparison with the CHA_2_DS_2_-VASc

The current tool used for the prediction of ischemic stroke occurrence among AF patients is CHA_2_DS_2_-VASc [56] which considers multiple risk factors; age, sex, heart failure, hypertension, stroke, vascular disease, diabetes. Thus, we decided to compare the performance of our best ML model, XGBoost (Table 4), with CHA_2_DS_2_-VASc in UKB. To construct the CHA_2_DS_2_-VASc we employed the codes described in *Supplementary_Material.xlsx (Table_S10)*. The AUROC and 95% CI for the CHA_2_DS_2_-VASc and XGBoost was 0.611 (0.585 – 0.638) and 0.631 (0.604 – 0.657) in the test set, respectively. The improved AUROC in the XGBoost model compared to CHA_2_DS_2_-VASc was statistically significant based on DeLong’s test for difference between the two models (pvalue=2.20E-06). Furthermore, SHAP analysis for the XGBoost model (Figure 4), shows that there is a significant number of peripheral blood markers associated with ischemic stroke, which are overlooked from CHA_2_DS_2_-VASc.

## Discussion

### *Comparison of the performance of ML models for prediction of AF or* ischemic stroke in patients with AF

We assessed six ML models in total for prediction of AF (XGBoost, LightGBM, RF, DNN, penalised LR) or ischemic stroke in patients with AF (XGBoost, LightGBM, RF, DNN, SVM, penalised LR) and employed SHAP analysis to rank features for predictive importance in a model. To the best of our knowledge, this is the first study using ML models to predict AF and ischemic stroke occurrence in AF patients in UKB. SHAP analysis was successful in the visualisation of non-linear relationships between the features used for prediction and the outcome. Additionally, the direction of the SHAP values for the top 20 features is in agreement with what has been reported so far in the literature. We found that the ensemble learning models LightGBM (best for AF prediction) and XGBoost (best for prediction of ischemic stroke in patients with AF) achieved higher AUROCs compared to the other examined models, suggesting that these models have better generalisation. In the case of models for AF prediction, DeLong’s test showed that penalised LR model had a lower AUROC compared to all other models and these differences were statistically significant (e.g., pvalue=1.38E-32 with LightGBM), indicating that ML models capture useful information by modeling non-linear associations. However, we cannot exclude that the performance of the examined ML models in this study may differ when applied to another dataset. For this reason, validation in datasets with different patient characteristics will be required in order to generalise these findings.

### AF results

Advancing age has been shown to be one of the most important risk factors for AF [3, 4, 57-60] which in our LightGBM model for AF prediction was ranked as the second most important feature. The third most important feature in our model was the AF SCORE, a set of 94 genome-wide variants associated with AF and explaining 42% of the heritability in Europeans [27], which as expected had a positive impact on the model output, i.e. the higher the AF score the higher the risk of developing AF. Thus, our results endorse the likely clinical utility of an AF score in disease prediction [61]. However, an optimised AF score for prediction in multi-ethnic populations such as the UK population will be required prior to considering clinical use. Interestingly, standing height was ranked as the fourth most significant feature in our best performing model for AF prediction. Greater height has been identified as a risk factor for AF in several studies [2, 9, 62-64] and in both males [10, 65] and females [65, 66]. Some studies report that taller people have greater heart chamber size [9, 10, 63, 65, 66], meaning a larger left atrial size, which may be potential explanation albeit not a very robust one as AF is driven by left atrial stretch and fibrosis. Two other anthropometric traits, weight and waist circumference, ranked just below standing height. Obesity is associated with increased risk of left atrial enlargement, atrial fibrosis, electrical derangements of the atria, impaired diastolic function, inflammation and accumulation of pericardial fat, which are all key mechanisms in the pathogenesis of AF [10, 62, 65, 66]. The ranking of sex as the 7^th^ most significant feature in the model is also in agreement with epidemiological studies reporting sex differences in AF, males are at higher risk which is in agreement with our results, along with the electrophysiologic properties of the atria and structural remodelling[59, 60]. Our analysis also found that participants with lower albumin levels (feature ranked 9^th^) had an increased risk of AF. This is in agreement with findings in two recent studies: Liao et.al. [67] found that albumin was inversely associated with risk of AF in the Atherosclerosis Risk in Communities (ARIC) study and the meta-analysis from Wang et al. [68] revealed that an increase in albumin level decreased the risk of AF. However, low albumin levels are associated with poor health overall and therefore we cannot exclude confounding. Among the remaining 20 most significant features in the model it is worth noting that (i) direct bilirubin (13/20) has been reported as an important independent risk factor for AF development in both thyrotoxic patients and a study in postoperative cardiac surgery[69, 70], (ii) urate (14/20) has been reported to increase the risk of AF [71] and be causally associated to AF through MR analysis in Koreans [72], and (iii) the positive effect of increased testosterone (17/20) on risk of AF has been reported in males but not in females in the ARIC study [73]. Finally, only two of the 20 top features have some conflicting data in the literature. FEV-1 levels (16/20) has an increased risk of AF as shown in other studies [74] but the Korean National Health and Nutritional Examination Survey reported an adverse association between FEV-1 and AF development [75] whereas triglycerides (20/20) contribute to increased risk of AF, but a study in Chinese participants showed no evidence of association between triglycerides and incidence of AF [76].

### AF & Ischemic stroke results

In our study, the XGBoost model was the best in predicting stroke in AF patients (AUROC 0.631) and showed that it performs better, albeit marginal this result was statistically significant (pvalue 2.20E-06 for DeLong’s test), than CHA_2_DS_2_-VASc. Unexpectedly, the genetic risk score for stroke (STROKE score [28]) was not among the top 20 features of our model although ischemic stroke is highly heritable [77, 78]. In the top 20 most significant features, medium to high feature values of HbA1c ranked third after sex and was associated with increased risk of stroke in AF patients. This is in agreement with the Clalit Health Services electronic medical records database from Israel, where participants with diabetes and AF were found to have an increased risk of stroke when their HbA1C levels were ranging from medium to high [79]. The fourth most significant feature was albumin which ranked 9^th^ in the AF prediction model suggesting a stronger relationship with ischemic stroke in AF patients than AF per se. A study in Japanese, has reported lower albumin levels to be associated with an increased risk of ischemic stroke in both sexes independently of AF status [80]. Four other blood biomarkers, creatinine, alkaline phosphatase, LDL cholesterol, and Lipoprotein A (Lp(a)) ranked among the top 20 features (6^th^,7^th^,10^th^, and 17^th^ respectively). These results are in agreement with the China National Stroke Registry reporting an association between high levels of alkaline phosphatase with recurrent stroke [81] and the Copenhagen General Population Study showing that high levels of Lp(a) were associated with increased risk of ischemic stroke [82, 83]. It is worth noting that the association of Lp(a) to increased risk of ischemic stroke although true for all examined ancestries it varies in strength e.g. higher in African than European Americans [84]. Interestingly, the use of creatinine as marker for increased risk of ischemic stroke in AF patients has not been previously reported and will merit further investigation. Lastly, the 20^th^ feature identified from the SHAP analysis – time spent watching television – could be considered as a surrogate marker for luck of sleep and physical inactivity. A study by Katzmarzyk et. al. [85], showed that physical inactivity increases the risk of stroke risk whereas a study in UKB, showed a dose-response joint association of sleep scores and physical inactivity with ischemic stroke mortality [86].

In summary, we present ML models for the prediction of AF and stroke in AF patients (XGBoost) respectively that have the potential for clinical use but validation in further independent studies is required. Importantly, the models will need to be validated across all ancestries as some features vary by ethnicity e.g., Lp(a) and AF genetic score. Our results endorse the incorporation of a number of routinely measured blood biomarkers whereas they support the inclusion of a genetic score only in the model for AF prediction.

## Data Availability

The data are from UK Biobank

## Footnotes

1 This research utilised Queen Mary’s Apocrita HPC facility, supported by QMUL Research-IT. http://doi.org/10.5281/zenodo.438045

## References

1. Khurshid, S., et al., Performance of Atrial Fibrillation Risk Prediction Models in Over 4 Million Individuals. Circ Arrhythm Electrophysiol, 2021. 14(1): p. e008997.

2. Allan, V., et al., Are cardiovascular risk factors also associated with the incidence of atrial fibrillation? A systematic review and field synopsis of 23 factors in 32 population-based cohorts of 20 million participants. Thromb Haemost, 2017. 117(5): p. 837–850.

3. Kornej, J., et al., Epidemiology of Atrial Fibrillation in the 21st Century: Novel Methods and New Insights. Circ Res, 2020. 127(1): p. 4–20.

4. Blum, S., et al., Incidence and predictors of atrial fibrillation progression: A systematic review and meta-analysis. Heart Rhythm, 2019. 16(4): p. 502–510.

5. Poorthuis, M.H.F., et al., Utility of risk prediction models to detect atrial fibrillation in screened participants. Eur J Prev Cardiol, 2021. 28(6): p. 586–595.

6. Benjamin, E.J., et al., Heart Disease and Stroke Statistics-2019 Update: A Report From the American Heart Association. Circulation, 2019. 139(10): p. e56–e528.

7. Chen, Y., et al., Classification of short single-lead electrocardiograms (ECGs) for atrial fibrillation detection using piecewise linear spline and XGBoost. Physiol Meas, 2018. 39(10): p. 104006.

8. Essa, H., A.M. Hill, and G.Y.H. Lip, Atrial Fibrillation and Stroke. Card Electrophysiol Clin, 2021. 13(1): p. 243–255.

9. Park, Y.M., et al., Height is associated with incident atrial fibrillation in a large Asian cohort. Int J Cardiol, 2020. 304: p. 82–84.

10. Andersen, K., et al., Body size and risk of atrial fibrillation: a cohort study of 1.1 million young men. J Intern Med, 2018. 283(4): p. 346–355.

11. Healey, J.S., G. Amit, and T.S. Field, Atrial fibrillation and stroke: how much atrial fibrillation is enough to cause a stroke? Curr Opin Neurol, 2020. 33(1): p. 17–23.

12. Joo, G., et al., Clinical Implication of Machine Learning in Predicting the Occurrence of Cardiovascular Disease Using Big Data (Nationwide Cohort Data in Korea). IEEE Access, 2020. 8: p. 157643–157653.

13. Tseng, P.Y., et al., Prediction of the development of acute kidney injury following cardiac surgery by machine learning. Crit Care, 2020. 24(1): p. 478.

14. Xu, Y., et al., Machine Learning Algorithms for Predicting the Recurrence of Stage IV Colorectal Cancer After Tumor Resection. Sci Rep, 2020. 10(1): p. 2519.

15. Qiu, H., et al., Machine learning approaches to predict peak demand days of cardiovascular admissions considering environmental exposure. BMC Med Inform Decis Mak, 2020. 20(1): p. 83.

16. Raghunath, S., et al., Deep Neural Networks Can Predict New-Onset Atrial Fibrillation From the 12-Lead ECG and Help Identify Those at Risk of Atrial Fibrillation-Related Stroke. Circulation, 2021. 143(13): p. 1287–1298.

17. Su, P.Y., et al., Machine Learning Models for Predicting Influential Factors of Early Outcomes in Acute Ischemic Stroke: Registry-Based Study. JMIR Med Inform, 2022. 10(3): p. e32508.

18. Chen, T. and C. Guestrin, XGBoost: A Scalable Tree Boosting System, in Proceedings of the 22nd ACM SIGKDD International Conference on Knowledge Discovery and Data Mining. 2016, Association for Computing Machinery: San Francisco, California, USA. p. 785–794.

19. Ke, G., et al., LightGBM: A Highly Efficient Gradient Boosting Decision Tree. 2017.

20. Breiman, L., Random forests. Machine learning, 2001. 45(1): p. 5–32.

21. Cortes, C. and V. Vapnik, Support-vector networks. Machine learning, 1995. 20(3): p. 273–297.

22. LeCun, Y., Y. Bengio, and G. Hinton, Deep learning. Nature, 2015. 521(7553): p. 436–44.

23. Tibshirani, R., Regression shrinkage and selection via the lasso. Journal of the Royal Statistical Society: Series B (Methodological), 1996. 58(1): p. 267–288.

24. Lundberg, S.M. and S.-I. Lee. A unified approach to interpreting model predictions. in Proceedings of the 31st international conference on neural information processing systems. 2017.

25. Millard, L.A.C., et al., Searching for the causal effects of body mass index in over 300 000 participants in UK Biobank, using Mendelian randomization. PLoS Genet, 2019. 15(2): p. e1007951.

26. Wu, P., et al., Mapping ICD-10 and ICD-10-CM Codes to Phecodes: Workflow Development and Initial Evaluation. JMIR Med Inform, 2019. 7(4): p. e14325.

27. Roselli, C., et al., Multi-ethnic genome-wide association study for atrial fibrillation. Nat Genet, 2018. 50(9): p. 1225–1233.

28. Malik, R., et al., Multiancestry genome-wide association study of 520,000 subjects identifies 32 loci associated with stroke and stroke subtypes. Nat Genet, 2018. 50(4): p. 524–537.

29. Pedregosa, F., et al., Scikit-learn: Machine learning in Python. the Journal of machine Learning research, 2011. 12: p. 2825–2830.

30. Lemaître, G., F. Nogueira, and C.K. Aridas, Imbalanced-learn: A python toolbox to tackle the curse of imbalanced datasets in machine learning. The Journal of Machine Learning Research, 2017. 18(1): p. 559–563.

31. Krawczyk, B., et al., Evolutionary undersampling boosting for imbalanced classification of breast cancer malignancy. Applied Soft Computing, 2016. 38: p. 714–726.

32. AlJame, M., et al., Ensemble learning model for diagnosing COVID-19 from routine blood tests. Inform Med Unlocked, 2020. 21: p. 100449.

33. Aridas, G.L.F.N.C.K., Imbalanced-learn: A Python Toolbox to Tackle the Curse of Imbalanced Datasets in Machine Learning. Journal of Machine Learning Research, 2017. 18(17): p. 1–5.

34. Berisha, V., et al., Digital medicine and the curse of dimensionality. NPJ Digit Med, 2021. 4(1): p. 153.

35. Ismael, R.-P., et al., When is resampling beneficial for feature selection with imbalanced wide data? Expert Systems with Applications, 2022. 188: p. 116015.

36. Kopitar, L., et al., Early detection of type 2 diabetes mellitus using machine learning-based prediction models. Sci Rep, 2020. 10(1): p. 11981.

37. Yu, S., et al., Exploring the relationship between 2D/3D landscape pattern and land surface temperature based on explainable eXtreme Gradient Boosting tree: A case study of Shanghai, China. Sci Total Environ, 2020. 725: p. 138229.

38. Chen, T., et al., Prediction of Extubation Failure for Intensive Care Unit Patients Using Light Gradient Boosting Machine. IEEE Access, 2019. 7: p. 150960–150968.

39. Zhang, D. and Y. Gong, The Comparison of LightGBM and XGBoost Coupling Factor Analysis and Prediagnosis of Acute Liver Failure. IEEE Access, 2020. 8: p. 220990–221003.

40. Rodriguez-Perez, R. and J. Bajorath, Interpretation of machine learning models using shapley values: application to compound potency and multi-target activity predictions. J Comput Aided Mol Des, 2020. 34(10): p. 1013–1026.

41. Bizopoulos, P. and D. Koutsouris, Deep Learning in Cardiology. IEEE Rev Biomed Eng, 2019. 12: p. 168–193.

42. Wainberg, M., et al., Deep learning in biomedicine. Nat Biotechnol, 2018. 36(9): p. 829–838.

43. Park, D.J., et al., Development of machine learning model for diagnostic disease prediction based on laboratory tests. Sci Rep, 2021. 11(1): p. 7567.

44. Zou, J., et al., A primer on deep learning in genomics. Nat Genet, 2019. 51(1): p. 12–18.

45. Futoma, J., J. Morris, and J. Lucas, A comparison of models for predicting early hospital readmissions. J Biomed Inform, 2015. 56: p. 229–38.

46. Zhao, Y., et al., Ensemble learning predicts multiple sclerosis disease course in the SUMMIT study. NPJ Digit Med, 2020. 3: p. 135.

47. Park, J.I., et al., Knowledge Discovery With Machine Learning for Hospital-Acquired Catheter-Associated Urinary Tract Infections. Comput Inform Nurs, 2020. 38(1): p. 28–35.

48. Unwin, N., et al., Body mass index, waist circumference, waist-hip ratio, and glucose intolerance in Chinese and Europid adults in Newcastle, UK. J Epidemiol Community Health, 1997. 51(2): p. 160–6.

49. Ballout, N., C. Garcia, and V. Viallon, Sparse estimation for case-control studies with multiple disease subtypes. Biostatistics, 2021. 22(4): p. 738–755.

50. Alharthi, A.M., et al., Quantitative structure-activity relationship model for classifying the diverse series of antifungal agents using ratio weighted penalized logistic regression. SAR QSAR Environ Res, 2020. 31(8): p. 571–583.

51. Lundberg, S.M., et al., From Local Explanations to Global Understanding with Explainable AI for Trees. Nat Mach Intell, 2020. 2(1): p. 56–67.

52. Rodriguez-Perez, R. and J. Bajorath, Interpretation of Compound Activity Predictions from Complex Machine Learning Models Using Local Approximations and Shapley Values. J Med Chem, 2020. 63(16): p. 8761–8777.

53. Lundberg, S.M., G.G. Erion, and S.-I. Lee Consistent Individualized Feature Attribution for Tree Ensembles. 2018. arXiv:1802.03888.

54. Molnar, C., Interpretable Machine Learning: A Guide for Making Black Box Models Explainable. Published online. 2019.

55. Van Rossum, G. and F.L. Drake, The python language reference manual. 2011:Network Theory Ltd.

56. Lip, G.Y., et al., Refining clinical risk stratification for predicting stroke and thromboembolism in atrial fibrillation using a novel risk factor-based approach: the euro heart survey on atrial fibrillation. Chest, 2010. 137(2): p. 263–72.

57. Cochet, H., et al., Predictors of future onset of atrial fibrillation in hypertrophic cardiomyopathy. Arch Cardiovasc Dis, 2018. 111(10): p. 591–600.

58. Chung, M.K., et al., Lifestyle and Risk Factor Modification for Reduction of Atrial Fibrillation: A Scientific Statement From the American Heart Association. Circulation, 2020. 141(16): p. e750–e772.

59. Westerman, S. and N. Wenger, Gender Differences in Atrial Fibrillation: A Review of Epidemiology, Management, and Outcomes. Curr Cardiol Rev, 2019. 15(2): p. 136–144.

60. Gerdts, E. and V. Regitz-Zagrosek, Sex differences in cardiometabolic disorders. Nat Med, 2019. 25(11): p. 1657–1666.

61. Lozano-Velasco, E., et al., Genetics and Epigenetics of Atrial Fibrillation. Int J Mol Sci, 2020. 21(16).

62. Feng, T., et al., Weight and weight change and risk of atrial fibrillation: the HUNT study. Eur Heart J, 2019. 40(34): p. 2859–2866.

63. Marott, J.L., et al., Increasing population height and risk of incident atrial fibrillation: the Copenhagen City Heart Study. Eur Heart J, 2018. 39(45): p. 4012–4019.

64. Sohail, H., et al., The height as an independent risk factor of atrial fibrillation: A review. Indian Heart J, 2021. 73(1): p. 22–25.

65. Johansson, C., et al., Weight, height, weight change, and risk of incident atrial fibrillation in middle-aged men and women. J Arrhythm, 2020. 36(6): p. 974–981.

66. Persson, C.E., et al., Young women, body size and risk of atrial fibrillation. Eur J Prev Cardiol, 2018. 25(2): p. 173–180.

67. Liao, L.Z., et al., Serum albumin and atrial fibrillation: insights from epidemiological and mendelian randomization studies. Eur J Epidemiol, 2020. 35(2): p. 113–122.

68. Wang, Y., et al., Relationship Between Serum Albumin and Risk of Atrial Fibrillation: A Dose-Response Meta-Analysis. Front Nutr, 2021. 8: p. 728353.

69. Sun, D., et al., Direct bilirubin level is an independent risk factor for atrial fibrillation in thyrotoxic patients receiving radioactive iodine therapy. Nucl Med Commun, 2019. 40(12): p. 1289–1294.

70. Turkkolu, S.T., E. Selcuk, and C. Koksal, Biochemical predictors of postoperative atrial fibrillation following cardiac surgery. BMC Cardiovasc Disord, 2021. 21(1): p. 167.

71. Zhang, J., et al., Serum uric acid and incident atrial fibrillation: A systematic review and doseresponse meta-analysis. Clin Exp Pharmacol Physiol, 2020. 47(11): p. 1774–1782.

72. Hong, M., et al., A mendelian randomization analysis: The causal association between serum uric acid and atrial fibrillation. Eur J Clin Invest, 2020. 50(10): p. e13300.

73. Berger, D., et al., Plasma total testosterone and risk of incident atrial fibrillation: The Atherosclerosis Risk in Communities (ARIC) study. Maturitas, 2019. 125: p. 5–10.

74. Au Yeung, S.L., et al., Impact of lung function on cardiovascular diseases and cardiovascular risk factors: a two sample bidirectional Mendelian randomisation study. Thorax, 2022. 77(2): p. 164–171.

75. Lee, S.N., et al., Association between lung function and the risk of atrial fibrillation in a nationwide population cohort study. Sci Rep, 2022. 12(1): p. 4007.

76. Li, X., et al., Lipid profile and incidence of atrial fibrillation: A prospective cohort study in China. Clin Cardiol, 2018. 41(3): p. 314–320.

77. O’Sullivan, J.W., et al., Combining Clinical and Polygenic Risk Improves Stroke Prediction Among Individuals With Atrial Fibrillation. Circ Genom Precis Med, 2021. 14(3): p. e003168.

78. Bevan, S., et al., Genetic heritability of ischemic stroke and the contribution of previously reported candidate gene and genomewide associations. Stroke, 2012. 43(12): p. 3161–7.

79. Kezerle, L., et al., Relation of Hemoglobin A1C Levels to Risk of Ischemic Stroke and Mortality in Patients With Diabetes Mellitus and Atrial Fibrillation. Am J Cardiol, 2022. 172: p. 48–53.

80. Li, J., et al., Serum Albumin and Risks of Stroke and Its Subtypes-The Circulatory Risk in Communities Study (CIRCS). Circ J, 2021. 85(4): p. 385–392.

81. Zong, L., et al., Alkaline Phosphatase and Outcomes in Patients With Preserved Renal Function: Results From China National Stroke Registry. Stroke, 2018. 49(5): p. 1176–1182.

82. Langsted, A., B.G. Nordestgaard, and P.R. Kamstrup, Elevated Lipoprotein(a) and Risk of Ischemic Stroke. J Am Coll Cardiol, 2019. 74(1): p. 54–66.

83. Kamstrup, P.R., Lipoprotein(a) and Cardiovascular Disease. Clin Chem, 2021. 67(1): p. 154–166.

84. Kumar, P., et al., Lipoprotein (a) level as a risk factor for stroke and its subtype: A systematic review and meta-analysis. Sci Rep, 2021. 11(1): p. 15660.

85. Katzmarzyk, P.T., et al., Physical inactivity and non-communicable disease burden in low-income, middle-income and high-income countries. Br J Sports Med, 2022. 56(2): p. 101–106.

86. Huang, B.H., et al., Sleep and physical activity in relation to all-cause, cardiovascular disease and cancer mortality risk. Br J Sports Med, 2022. 56(13): p. 718–724.

